# Toward Using Twitter Data to Monitor Covid-19 Vaccine Safety in Pregnancy

**DOI:** 10.1101/2021.09.15.21263653

**Authors:** Ari Z. Klein, Karen O’Connor, Graciela Gonzalez-Hernandez

## Abstract

**Background:** Coronavirus Disease 2019 (Covid-19) during pregnancy is associated with an increased risk of maternal death, intensive care unit (ICU) admission, and preterm birth; however, many people who are pregnant refuse to receive Covid-19 vaccination because of a lack of safety data.

**Objective:** The objective of this preliminary study was to assess whether we could identify (1) users who have reported on Twitter that they received Covid-19 vaccination during pregnancy or the periconception period, and (2) reports of their pregnancy outcomes.

**Methods:** We searched for reports of Covid-19 vaccination in a large collection of tweets posted by users who have announced their pregnancy on Twitter. To help determine if users were vaccinated during pregnancy, we drew upon a natural language processing (NLP) tool that estimates the timeframe of the prenatal period. For users who posted tweets with a timestamp indicating they were vaccinated during pregnancy, we drew upon additional NLP tools to help identify tweets that report their pregnancy outcomes.

**Results:** Upon manually verifying the content of tweets detected automatically, we identified 150 users who reported on Twitter that they received at least one dose of Covid-19 vaccination during pregnancy or the periconception period. Among the 60 completed pregnancies, we manually verified at least one reported outcome for 45 (75%) of them.

**Conclusions:** Given the limited availability of data on Covid-19 vaccine safety in pregnancy, Twitter can be a complementary resource for potentially increasing the acceptance of Covid-19 vaccination in pregnant populations. Directions for future work include developing machine learning algorithms to detect a larger number of users for observational studies.

## Introduction

Coronavirus Disease 2019 (Covid-19) during pregnancy is associated with an increased risk of maternal death, intensive care unit (ICU) admission, and preterm birth [1]; however, in the United States, receipt of Covid-19 vaccination during pregnancy is low [2]. Surveys indicated that the most common reason for refusing Covid-19 vaccination during pregnancy was a lack of safety data [3], which are limited because people who were pregnant were excluded from preauthorization clinical trials. The Centers for Disease Control and Prevention (CDC) recently released the first United States data on Covid-19 vaccine safety in pregnancy, based on post-vaccination health information reported by participants voluntarily enrolled in *v-safe* [4]. According to the CDC, the preliminary data do not indicate any obvious safety signals, but continued monitoring is needed. The CDC also noted several potential limitations of their participant-reported data, including selection bias, reporting bias, misreporting, small sample size, and limited information on other risk factors.

Given the preliminary nature and potential limitations of the *v-safe* data, other sources of data should be explored in order to provide additional evidence that Covid-19 vaccination during pregnancy is safe. In the United States, 42% of people aged 18-29 and 27% of people aged 30-49 use Twitter [5], and our prior work [6] demonstrates that Twitter data can be used to assess outcomes associated with medication taken during pregnancy. Therefore, we hypothesized that Twitter also could be a source of data for monitoring the safety of Covid-19 vaccination received during pregnancy. While user-generated Twitter data may be subject to limitations similar to the *v-safe* data, the current availability of other sources of data is very limited. The objective of this preliminary study was to assess whether we could identify (1) users who have reported on Twitter that they received Covid-19 vaccination during pregnancy or the periconception period (within 30 days before their last menstrual period), and (2) reports of their pregnancy outcomes. As a complementary source of safety data, Twitter could help increase the acceptance of Covid-19 vaccination in pregnant populations.

## Methods

As a preliminary approach to detecting self-reports of Covid-19 vaccination on Twitter, we deployed six handwritten, high-precision regular expressions—search patterns that automatically match text strings—on a large collection of public tweets posted by users who have announced their pregnancy on Twitter [7]. To help determine if users were vaccinated during pregnancy or the periconception period, we used the timestamp of the tweets that matched the regular expressions and drew upon an automated natural language processing (NLP) tool that estimates the timeframe of the prenatal period based on tweets that report the baby’s gestational age, due date, or date of birth [8]. For users who posted tweets with a timestamp indicating they were vaccinated during pregnancy or the periconception period, we drew upon additional automated NLP tools to help identify tweets that report their adverse pregnancy outcomes, including miscarriage, stillbirth, preterm birth, low birthweight, birth defects, and neonatal intensive care unit admission [9-11]. To reduce the potential reporting bias in assuming that the lack of tweets self-reporting an adverse pregnancy outcome represents the lack of an adverse outcome, we also deployed an automated NLP tool to help identify reports that the baby was born at a gestational age of at least 37 weeks and a weight of at least 5 pounds and 8 ounces [12]. Tweets that report a gestational age of at least 37 weeks indicate the lack of miscarriage or preterm birth. Tweets that report a birthweight of at least 5 pounds and 8 ounces indicate the lack of low birthweight or, as a report of live birth, miscarriage or stillbirth.

## Results

Upon manually verifying the content of tweets detected automatically, we identified 150 users who reported on Twitter that they received at least one dose of Covid-19 vaccination during pregnancy or the periconception period. Table 1 presents examples of tweets that we used to identify these 150 users. In Table 1, User 1 reported being 16 weeks pregnant on June 15, 2021, so our automated tool [8] estimated that pregnancy began on February 23, 2021. User 1 reported receiving Covid-19 vaccination on March 24, 2021—approximately one month into pregnancy. User 2 reported being 13 weeks pregnant on June 21, 2021, so our automated tool [8] estimated that pregnancy began on March 22, 2021. User 2 reported receiving Covid-19 vaccination on March 6, 2021—during the periconception period. The tweets in Table 1 also show that some users report the vaccine manufacturer (e.g., *#PfizerVaccine*) or dose number (e.g., *second vaccine*), which can help distinguish mRNA vaccines from other types. Based on our estimates of the prenatal period for these 150 users, 90 (60%) of their pregnancies may have been ongoing. Among the 60 completed pregnancies, we manually verified at least one reported outcome for 45 (75%) of them. Table 2 presents the outcomes reported by these 45 users.

**Table 1.**
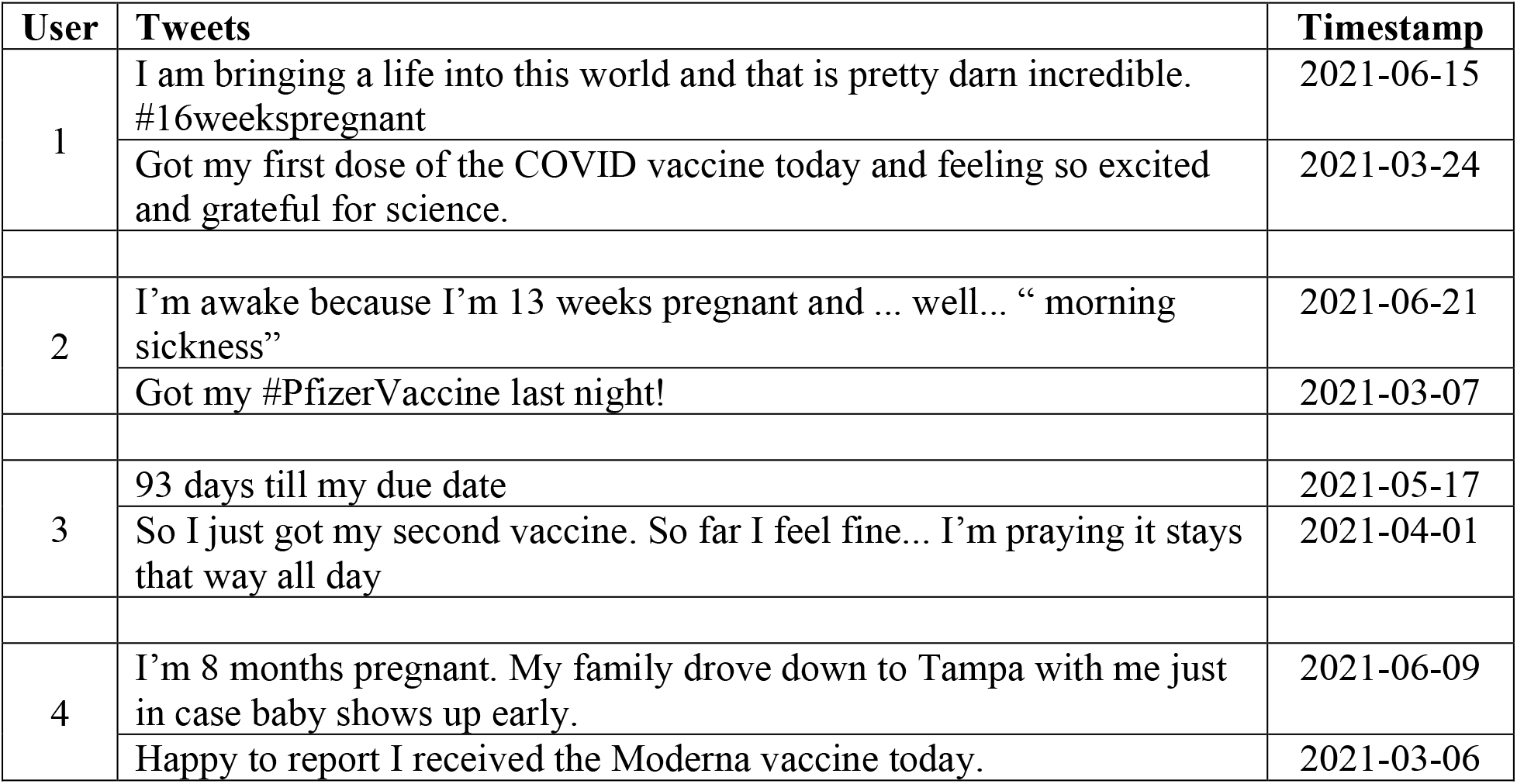
Sample tweets indicating that COVID-vaccination was received during pregnancy or the periconception period.

| User | Tweets | Timestamp |
| --- | --- | --- |
| 1 | I am bringing a life into this world and that is pretty darn incredible.<br>#16weekspregnant | 2021-06-15 |
|  | Got my first dose of the COVID vaccine today and feeling so excited and grateful for science. | 2021-03-24 |
| 2 | I'm awake because I'm 13 weeks pregnant and ... well... "morning sickness" | 2021-06-21 |
|  | Got my #PfizerVaccine last night! | 2021-03-07 |
| 3 | 93 days till my due date | 2021-05-17 |
|  | So I just got my second vaccine. So far I feel fine... I'm praying it stays that way all day | 2021-04-01 |
| 4 | I'm 8 months pregnant. My family drove down to Tampa with me just in case baby shows up early. | 2021-06-09 |
|  | Happy to report I received the Moderna vaccine today. | 2021-03-06 |

**Table 2.** Self-reported pregnancy outcomes for Twitter users who received COVID-19 vaccination during pregnancy or the periconception period.

| Self-Reported Outcome | No. (%) (n=45)<sup>a</sup> | Sample Tweet |
| --- | --- | --- |
| **Adverse outcomes** |  |  |
| Neonatal intensive care unit | 5 (11.1%) | I made a small human. So that's pretty cool. Now for a few weeks of NICU time. |
| Preterm birth (< 37 weeks) | 4 (8.9%) | She was born Jan. 11th...3 months early...stayed in the hospital until about 2.5 weeks ago... |
| Low birthweight (< 5 lbs 8 oz) | 1 (2.2%) | He weighed 3 lbs 9 ounces @ birth & we didn't have 1 thing that came close to fitting him. |
| Miscarriage | 1 (2.2%) | In the last 4 weeks, I've had a miscarriage... family death.... pet death... my car broke down... finals... |
| Stillbirth | 0 (0.0%) | -- |
| Birth defect | 0 (0.0%) | -- |

**Table 2 (cont'd).** Self-reported pregnancy outcomes for Twitter users who received COVID-19 vaccination during pregnancy or the periconception period.
| **Normal outcomes** |  |  |
| --- | --- | --- |
| Term ($\geq 37$ weeks)<sup>b</sup> | 39 (86.7%) | He made his debut at #37weeks. We got to the hospital by 6:15am, fully dilated by 7:45am, and he was here at 8:22am! |
| Normal birthweight ($\geq 5$ lbs 8 oz) | 7 (15.6%) | He arrived via c/section last night at 8:49pm. He was 7 lbs 11 oz. I can't believe he's mine! |

<sup>a</sup> Multiple outcomes were identified for some pregnancies, so the sum and percentage of the total outcomes are greater than n=45 and 100%, respectively.
<sup>b</sup> Pregnancies were included for which we did not find subsequent tweets explicitly indicating live birth.

## Discussion

The CDC states that continued monitoring is needed to further assess outcomes associated with Covid-19 vaccination during pregnancy, especially in early pregnancy and the periconception period [4], and suggests that additional evidence of Covid-19 vaccine safety in pregnancy is critical for increasing the acceptance of Covid-19 vaccination in pregnant populations [2]. Our study demonstrates that there are users who report on Twitter that they were vaccinated during pregnancy, including in early pregnancy, and the periconception period, and that many of them report their pregnancy outcomes. Although not directly comparable, out of the total number of pregnancies with a reported gestational age of at least 20 weeks, the proportion of preterm births reported on Twitter (9.09%) is similar to both the incidence in the United States prior to the Covid-19 pandemic (10.23%) [13] and the proportion reported by *v-safe* participants (9.4%) [4]. Given our initial small sample of Twitter users, it is not surprising that we did not detect any reports of birth defects or stillbirth, which have an incidence in the United States of 3% [14] and less than 1% [15], respectively. Nonetheless, our prior work [9-11] demonstrates that users do report these rare outcomes on Twitter. The small number of users identified in this study likely reflects that our regular expressions were limited in number and highly precise in order to facilitate a preliminary assessment of Covid-19 vaccination reports on Twitter.

## Conclusions

Given the limited availability of data on Covid-19 vaccine safety in pregnancy, Twitter can be a complementary resource for continued monitoring and potentially increasing the acceptance of Covid-19 vaccination in pregnant populations. Directions for future work include developing machine learning algorithms to detect a larger number of users who have reported on Twitter that they received Covid-19 vaccination during pregnancy or the periconception period, and conducting an observational study comparing their pregnancy outcomes to those of users who have announced their pregnancy on Twitter [7] but gave birth prior to the availability of Covid-19 vaccines.

## Data Availability

The Twitter data that was analyzed to identify pregnancy, Covid-19 vaccination, and pregnancy outcomes will be made available upon request.

## Acknowledgments

AZK developed the regular expressions, analyzed the Twitter data for pregnancy outcomes, and wrote the manuscript. KO analyzed the Twitter data to identify users who received Covid-19 vaccination during pregnancy, and edited the manuscript. GGH guided the overall study design and edited the manuscript. This work was supported by the National Institutes of Health (NIH) National Library of Medicine (NLM; grant number R01LM011176).

## References

1. Allotey J, Stallings E, Bonet M, Yap M, Chatterjee S, Kew T, Debenham L, Llavall AC, Dixit A, Zhou D, Balaji R, Lee SI, Qiu X, Yuan M, Coomar D, Sheikh J, Lawson H, Ansari K, van Wely M, van Leeuwen E, Kostova E, Kunst H, Khalil A, Tiberi S, Brizuela V, Broutet N, Kara E, Kim CR, Thorson A, Oladapo OT, Mofenson L, Zamora J, Thangaratinam S, PregCOV-19 Living Systematic Review Consortium. Clinical manifestations, risk factors, and maternal and perinatal outcomes of coronavirus disease 2019 in pregnancy: living systematic review and meta-analysis. BMJ 2020;370:m3320.

2. Razzaghi H, Meghani M, Pingali C, Crane B, Naleway A, Weintraub C, Kenigsberg TA, Lamias MJ, Irving SA, Kauffman TL, Vesco KK, Daley MF, DeSilva M, Donahue J, Getahun D, Glenn S, Hambidge SJ, Jackson L, Lipkind HS, Nelson J, Zerbo O, Oduyebo T, Singleton JA, Patel SA. COVID-19 vaccination coverage among pregnant women during pregnancy - eight integrated health care organizations, United States, December 14, 2020-May 8, 2021. MMWR Morb Mortal Wkly Rep 2021;70(24):895–899.

3. Ayhan SG, Oluklu D, Atalay A, Beser DM, Tanacan A, Tekin OM, Sahin D. COVID-19 vaccine acceptance in pregnant women. Int J Gynaecol Obstet 2021;154(2):291–296.

4. Shimabukuro TT, Kim SY, Myers TR, Moro PL, Oduyebo T, Panagiotakopoulos L, Marquez PL, Olson CK, Liu R, Chang KT, Ellington SR, Burkel VR, Smoots AN, Green CJ, Licata C, Zhang BC, Alimchandani M, Mba-Jonas A, Martin SW, Gee JM, Meaney-Delman DM, CDC v-safe COVID-19 Pregnancy Registry Team. Preliminary findings of mRNA Covid-19 vaccine safety in pregnant persons. N Engl J Med 2021;384(24):2273–2282.

5. Auxier B, Anderson M. Social media use in 2021. 2021 April 7. URL: https://www.pewresearch.org/internet/2021/04/07/social-media-use-in-2021/ [accessed 2021-08-02]

6. Golder S, Chiuve S, Weissenbacher D, Klein A, O’Connor K, Bland M, Malin M, Bhattacharya M, Scarazzini LJ, Gonzalez-Hernandez G. Pharmacoepidemiologic evaluation of birth defects from health-related postings in social media during pregnancy. Drug Saf 2019;42(3):389–400.

7. Sarker A, Chandrashekar P, Magge A, Cai H, Klein A, Gonzalez G. Discovering cohorts of pregnant women from social media for safety surveillance and analysis. J Med Internet Res 2017;19(10):e361.

8. Rouhizadeh M, Magge A, Klein A, Sarker A, Gonzalez G. A rule-based approach to determining pregnancy timeframe from contextual social media postings. 2018 Presented at: 2018 International Conference on Digital Health; April 23-26, 2018; Lyon, France p. 16–20.

9. Klein AZ, Sarker A, Cai H, Weissenbacher D, Gonzalez-Hernandez G. Social media mining for birth defects research: a rule-based, bootstrapping approach to collecting data for rare health-related events on Twitter. J Biomed Inform 2018;87:68–78.

10. Klein AZ, Sarker A, Weissenbacher D, Gonzalez-Hernandez G. Towards scaling Twitter for digital epidemiology of birth defects. NPJ Digit Med 2019;2:96.

11. Klein AZ, Cai H, Weissenbacher D, Gonzalez-Hernandez G. A natural language processing pipeline to advance the use of Twitter data for digital epidemiology of adverse pregnancy outcomes. J Biomed Inform 2020;112S:100076.

12. Klein AZ, Gebreyesus A, Gonzalez-Hernandez G. Automatically identifying comparator groups on Twitter for digital epidemiology of pregnancy outcomes. AMIA Jt Summits Transl Sci Proc 2020:317–325.

13. Martin JA, Hamilton BE, Osterman MJK, Driscoll AK. Births: final data for 2019. Natl Vital Stat Rep 2021;70(2):1–51.

14. Centers for Disease Control and Prevention. Update on overall prevalence of major birth defects—Atlanta, Georgia, 1978-2005. MMWR Morb Mortal Wkly Rep 2008;57(1):1–5.

15. Hoyert DL, Gregory ECW. Cause-of-death data from the fetal death file, 2015-2017. Natl Vital Stat Rep 2020;69(4):1–20.

